# A Clinical Consensus Treatment Algorithm for Patients with High-tone Pelvic Floor Dysfunction: a Multidisciplinary Delphi Study

**DOI:** 10.1101/2023.08.11.23293953

**Authors:** Michele Torosis, Erin Carey, Kristin Christensen, Melissa R. Kaufman, Kimberly Kenton, Rhonda Kotarinos, H. Henry Lai, Una Lee, Jerry L. Lowder, Melanie Meister, Theresa Spitznagle, Kelly Wright, A. Lenore Ackerman

## Abstract

**BACKGROUND:** High-tone pelvic floor dysfunction (HTPFD) is a neuromuscular disorder of the pelvic floor characterized by non-relaxing pelvic floor muscles, resulting in lower urinary tract and defecatory symptoms, sexual dysfunction, and pelvic pain. Despite affecting 80% of women with chronic pelvic pain, there are no uniformly accepted guidelines to direct the management of these patients. We sought to develop evidence-and consensus-based clinical practice guidelines for management of HTPFD.

**METHODS AND FINDINGS:** A Delphi method of consensus development was used, comprising three survey rounds administered anonymously via web-based platform (Qualtrics® XM) to national experts in the field of HTPFD recruited through targeted invitation between September and December 2021. Twelve experts participated with backgrounds in urology, urogynecology, minimally invasive gynecology, and pelvic floor physical therapy (PFPT) participated. Panelists were asked to rate their agreement with rated evidence-based statements regarding HTPFD treatment. Statements reaching consensus were used to generate a consensus treatment algorithm. A total of 31 statements were reviewed by group members at the first Delphi round with 10 statements reaching consensus. 28 statements were reposed in the second round with 17 reaching consensus. The putative algorithm met clinical consensus in the third round. There was universal agreement for PFPT as first-line treatment for HTPFD. If satisfactory symptom improvement is reached with PFPT, the patient can be discharged with a home exercise program. If no improvement after PFPT, second line options include trigger or tender point injections, vaginal muscle relaxants, and cognitive behavioral therapy, all of which can also be used in conjunction with PFPT. Onabotulinumtoxin A injections should be used as third line with symptom assessment after 2-4two to four weeks. There was universal agreement that sacral neuromodulation is fourth line intervention. The largest identified barrier to care for these patients is access to PFPT. For patients who cannot access PFPT, experts recommend at-home, guided pelvic floor relaxation, self-massage with vaginal wands, and virtual PFPT visits.

**CONCLUSIONS:** A stepwise approach to the treatment of HTPFD is recommended, with patients often necessitating multiple lines of treatment either sequentially or in conjunction. However, PFPT should be offered first line.

**AUTHOR SUMMARY:** *Why was this study done?:* High tone pelvic floor dysfunction is a highly prevalent neuromuscular disorders causing a range of lower urinary tract and defecatory symptoms, sexual dysfunction, and pelvic pain. Currently, there is no consensus or accepted clinical guidance to direct the management of these patients.

*What did the researchers do and find?:* Experts in urology, urogynecology, minimally invasive gynecology, and pelvic floor physical therapy generated a set of consensus practice statements to help guide the management of HTPFD using a rigorous Delphi process. Four tiers of treatment, with pelvic floor physical therapy as the first line treatment, followed by vaginal muscle relaxants, tender point injections, and/or cognitive behavioral therapy as second line, pelvic floor trigger point injection as third-line, and sacral neuromodulation as fourth line, were recommended for HTPFD management. Exceptions to sequential progression through this algorithm may be needed for patients with limited regional or financial access to the recommended treatments.

*What do these findings mean?:* A clinical care pathway for high-tone pelvic floor dysfunction will enable more effective care of patients and empower future studies to measure the effectiveness of each treatment option in a more systematic fashion.

## INTRODUCTION

Chronic pelvic pain (CPP) is estimated to affect one quarter of women and cost the US healthcare system more than $5.8 billion annually. (1) (2) High-tone pelvic floor disorder (HTPFD), a chronic pelvic floor disorder characterized by tight, weakened, and/or painful pelvic floor muscles, is present in 60 to 90% of women with CPP. (1) (3) (4) Tension within the muscle interferes with its dynamic physiologic action, preventing appropriate coordination, contraction, and relaxation. This may be associated with a wide range of genitourinary complaints, including discomfort, lower urinary tract symptoms (LUTS), defecatory dysfunction, and dyspareunia. HTPFD can be the sole cause for pain and genitourinary complaints or can co-exist with other pelvic pain disorders, such as vulvodynia or endometriosis; however, it frequently goes both unrecognized and untreated, contributing to the poor outcomes often seen in these conditions.

Recognition and diagnosis of HTPFD is complicated by its wide spectrum of clinical presentations and symptomatic overlap with other gynecologic, urologic and colorectal syndromes. While chronic pain is a common presentation, many patients with HTPFD may seek medical care for LUTS or defecatory dysfunction as their primary complaint. A growing body of literature indicates that increased pelvic floor tone underlies many urinary, gastrointestinal, and sexual complaints, even in the absence of pain. (5) The etiology of HTPFD is equally broad, ranging from idiopathic, potentially incited by poor toileting habits or triggered by visceral dysfunction, including endometriosis or interstitial cystitis/bladder pain syndrome (IC/BPS), or musculoskeletal injuries, such as sacroiliac joint dysfunction or hip osteoarthritis, all highly prevalent conditions. Clinical identification of increased pelvic floor tone is subjective and provider-dependent, relying on discriminate vulvar, vaginal, and rectal examination to identify hypertonicity of the levator ani and obturator internus musculature. (6) (7)

Beyond the diagnosis, there is little guidance on treatment pathways. While multiple clinical trials and observational studies have sought to establish the efficacies of a range of treatments for HTPFD, there is considerable variability in the level of peer-reviewed evidence. In addition, head-to-head studies of therapeutic approaches are lacking, making it challenging for many providers to determine an effective course of treatment, particularly when access to certain therapeutic modalities, such as pelvic floor physical therapy (PFPT), can be limited. This lack of guidance often leaves patients unable to access effective treatment or make informed decisions surrounding their care.

In the absence of guidelines or strong level I evidence, we sought to generate a treatment algorithm based on using the Delphi method, a formal, systematic qualitative methodology, (8) to compile expert opinion statements supported by the available literature pertinent to the treatment of HTPFD. Compilation of these consensus statements into the first proposed comprehensive treatment algorithm for women with HTPFD sought to facilitate clinician’s efforts to provide the best treatment options for these frequently debilitated patients.

## METHODS

### Design and Participants

The HTPFD treatment consensus was developed using a modified Delphi process. After exemption from Institutional Review Board approval (UCLA IRB#21-001399), we identified a range of practitioners with expertise in the diagnosis and management of HTPFD. Ten to 15 experts have been described as a minimum number for Delphi methodology to yield sufficient results and ensure validity. (9) (10) Initial screening identified 32 practitioners who were either academic clinicians with a track record of publications or academic speaking on HTPFD and/or experts who had published online, book, and social media informational materials on HTPFD. Twenty-five of these practitioners confirmed that HTPFD currently comprised at least 20% of their patient population; 12 of these specialists agreed to participate, reflecting geographic diversity and balanced representation of the multiple specialties.

Prior to initiating the Delphi process, the intended scope of the practice algorithm was outlined, and group members were asked to review a panel of literature containing pertinent clinical trials and systematic reviews. The definition of HTPFD was provided to the participants as per the International Continence Society. (11) Web-based software Qualtrics (Provo, UT) was used to administer confidential surveys to participants.

### Delphi Study

The Delphi method, an iterative process that uses a systematic progression of repeated rounds of voting, is an effective process for determining expert group consensus where there is little definitive evidence. (8) The process was conducted in three phases which took place between September and December 2021 (**Figure 1**). To develop the initial statements, a literature review was conducted to identify current evidence regarding options and efficacy of treatments for HTPFD. Relevant information was collected from studies regarding best practices on how to treat HTPFD, which were formulated by the study team into 31 statements for the Round 1 questionnaire. Survey questions used a 5-point Likert scale for participants to rate their level of agreement with questions. (12) There was also opportunity for short comments explaining their rationale for agreement or disagreement with statement. Consensus was defined as 70% or more of the members voting in agreement. (13) (14) Statements with ≥ 70% of agreement (either strongly agree and agree; or strongly disagree and disagree) were interpreted as “appropriate” or “inappropriate” to include in the guidelines, respectively. Statements that did not meet consensus were revised and reposed to the group in the next round. New concepts that were presented by at least three individual experts were incorporated into the next round of statements.

**Figure 1.**
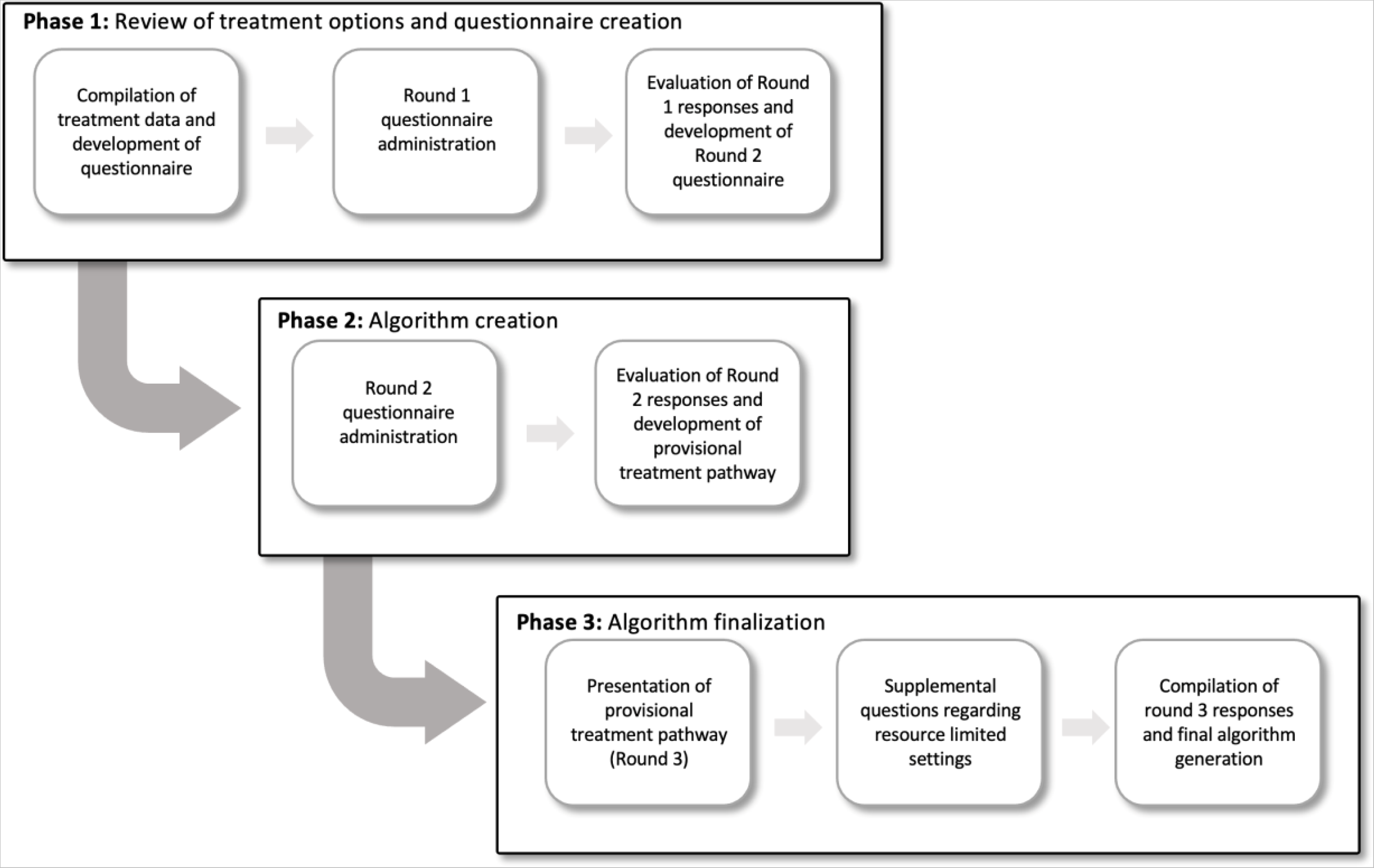
Diagrammatic Overview of Modified Delphi Process. Multiple questionnaire rounds were sent to the panel of experts to work toward a consensus opinion. The participants were allowed additional free-text responses to help generate revised statements in subsequent rounds. In later rounds, information brought forth by other experts was also included to help refine consensus statements. After the third round, a final treatment algorithm was generated from the consensus statements and approved by all members.

In phase two, the revised Round 2 questionnaire was sent to all participants and again participants were asked to rate level of agreement on a 5-point Likert scale. Results from Round 2 were analyzed, and a treatment pathway was developed by the study team based on consensus statements (Round 3). In phase three, the provisional treatment pathway was presented to experts and qualitative and quantitative agreement was captured. Experts were also asked additional qualitative questions about how they advise patients in resource limited settings.

### Statistical analysis

Aggregated data were stored and analyzed in Microsoft Excel for Mac (V16.16.16; Microsoft Corporation, Redmond, WA).

## RESULTS

A total of 4 American Board of Urology (ABU)-certified female pelvic medicine and reconstructive surgery (FPMRS) urologists, 3 American Board of Obstetrics and Gynecology (ABOG)-certified urogynecologists, 2 minimally invasive gynecologists, and 3 American Board of Physical Therapy Specialties (ABPTS)-certified Women’s Clinical Health Specialist physical therapists participated in this effort. The treatment consensus was developed and confirmed in an electronic, three-phase, modified Delphi process. A total of 31 statements were reviewed by the HTPFD expert group members at the first Delphi round with 10 statements reaching consensus. The 21 statements that did not reach consensus were reviewed and revised to clarify ambiguities. Some statements were divided into two statements to aid in clarification. In total, 28 statements were included in the second Delphi survey and 17 of these statements reached consensus (**Table 1**). After the second round, it was determined that the statements that had not reached consensus were because of absence of evidence rather than ambiguity in wording (**Appendix 1**). A treatment pathway (**Figure 2**) was generated and evaluated by the group members in the third Delphi survey, which met clinical consensus.

**Figure 2.**
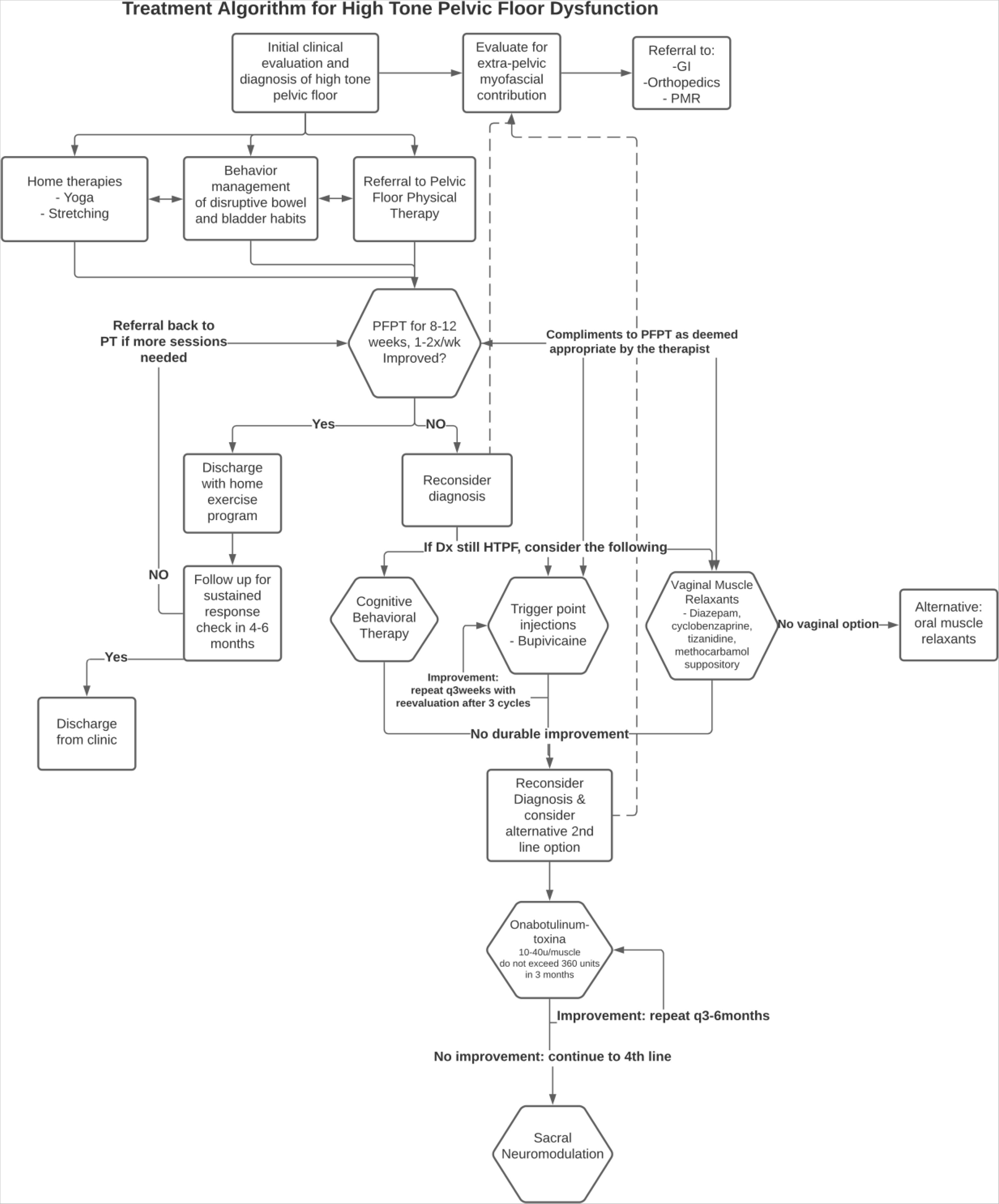
Treatment Algorithm for High Tone Pelvic Floor Dysfunction. The consensus statement regarding treatment approaches for HTPFD were organized into an algorithm with levels of treatment ranging from least invasive and most evidence-based to more invasive approaches with weaker supporting evidence.

**Table 1.**
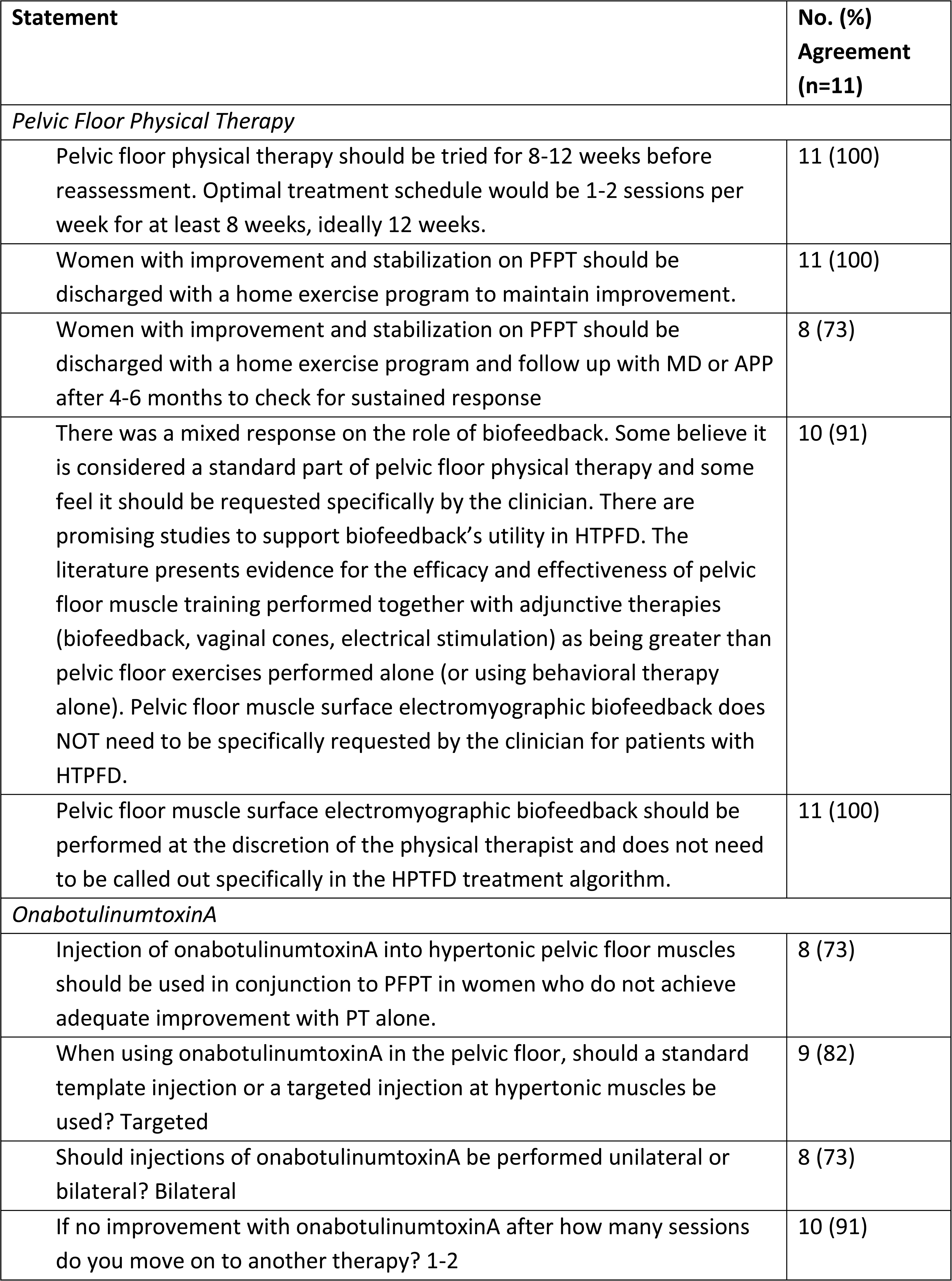
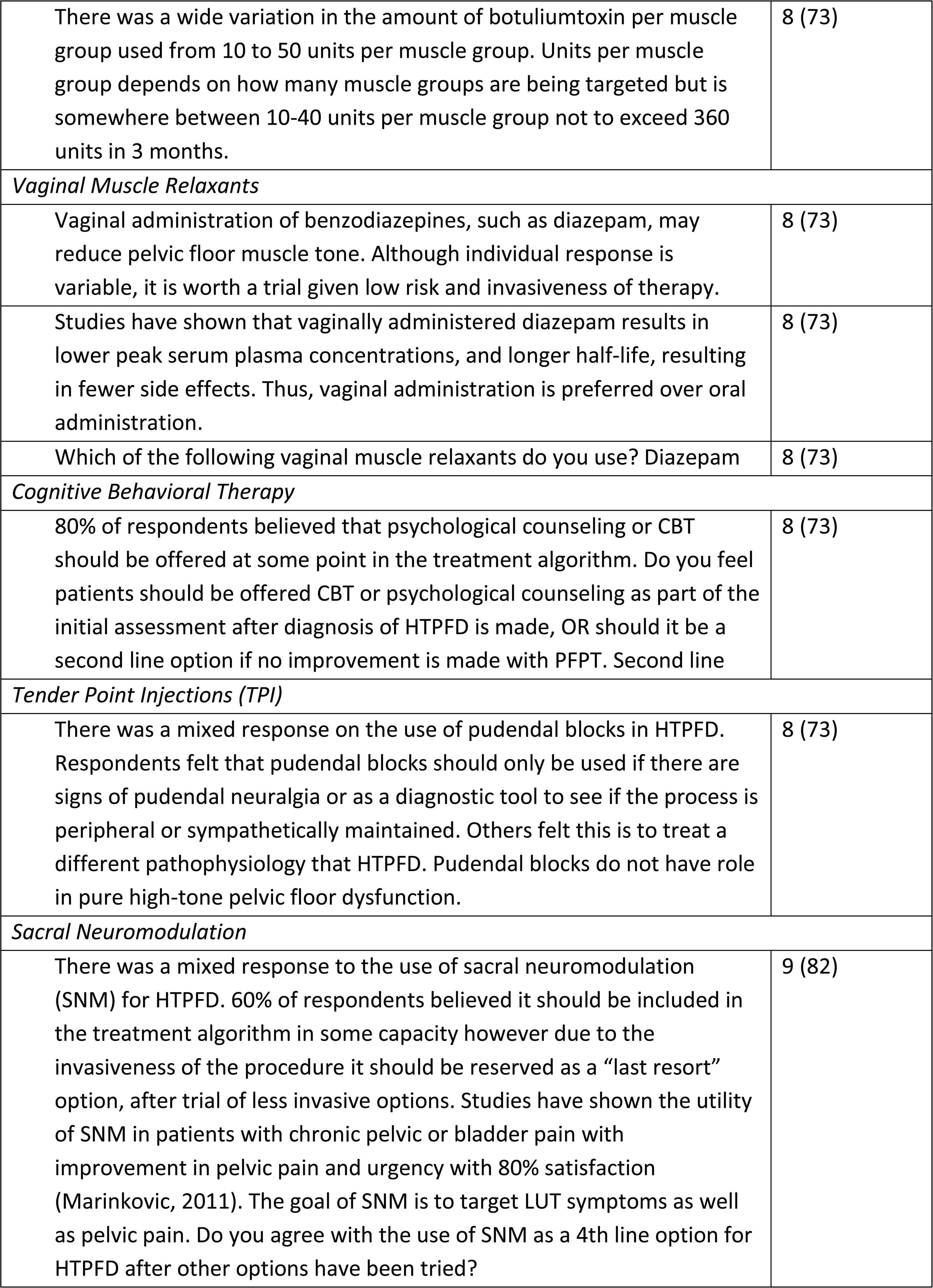
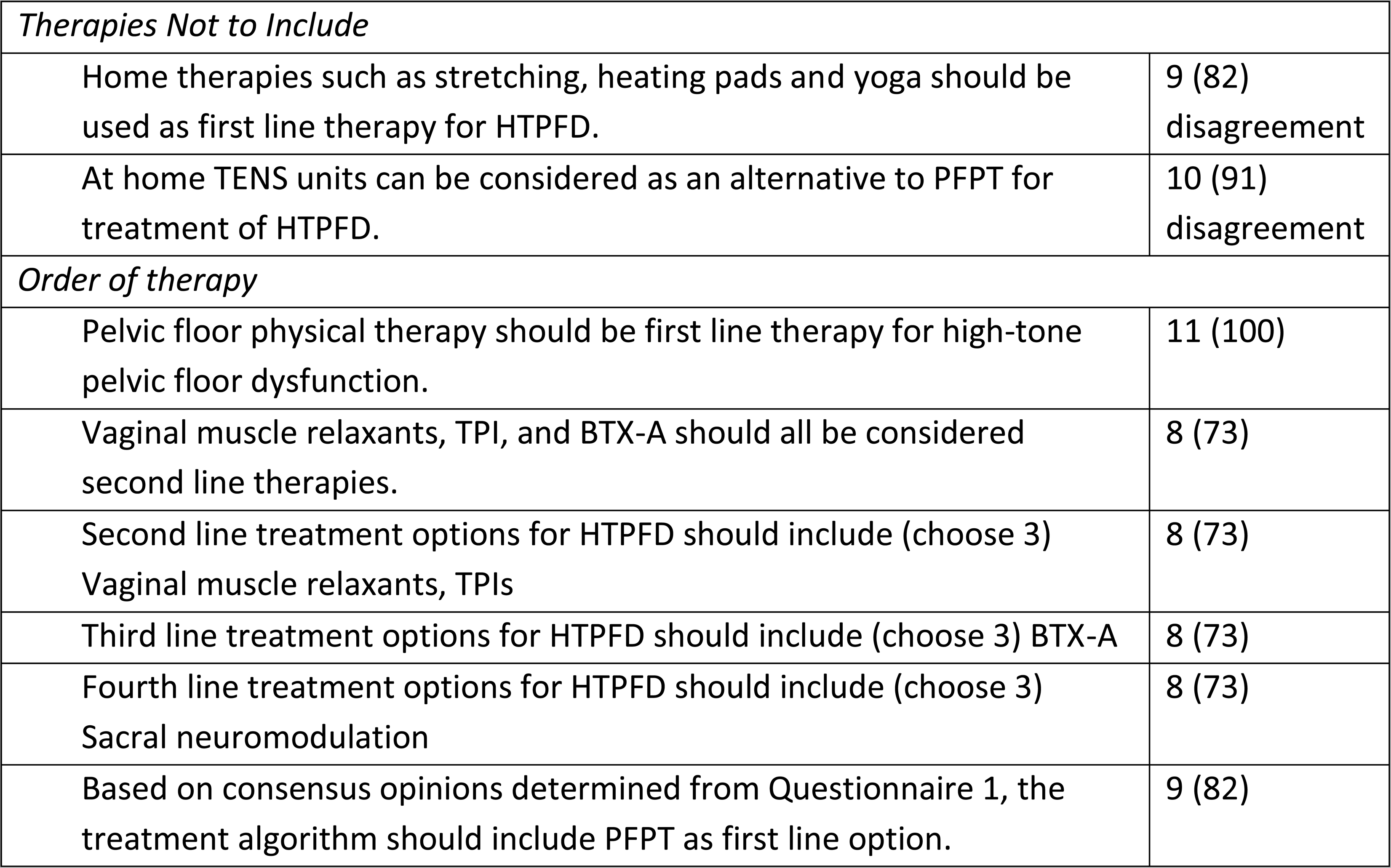
Statements That Met Consensus.

| Statement | No. (%) Agreement (n=11) |
| --- | --- |
| *Pelvic Floor Physical Therapy* |  |
| Pelvic floor physical therapy should be tried for 8-12 weeks before reassessment. Optimal treatment schedule would be 1-2 sessions per week for at least 8 weeks, ideally 12 weeks. | 11 (100) |
| Women with improvement and stabilization on PFPT should be discharged with a home exercise program to maintain improvement. | 11 (100) |
| Women with improvement and stabilization on PFPT should be discharged with a home exercise program and follow up with MD or APP after 4-6 months to check for sustained response | 8 (73) |
| There was a mixed response on the role of biofeedback. Some believe it is considered a standard part of pelvic floor physical therapy and some feel it should be requested specifically by the clinician. There are promising studies to support biofeedback's utility in HTPFD. The literature presents evidence for the efficacy and effectiveness of pelvic floor muscle training performed together with adjunctive therapies (biofeedback, vaginal cones, electrical stimulation) as being greater than pelvic floor exercises performed alone (or using behavioral therapy alone). Pelvic floor muscle surface electromyographic biofeedback does NOT need to be specifically requested by the clinician for patients with HTPFD. | 10 (91) |
| Pelvic floor muscle surface electromyographic biofeedback should be performed at the discretion of the physical therapist and does not need to be called out specifically in the HPTFD treatment algorithm. | 11 (100) |
| *OnabotulinumtoxinA* |  |
| Injection of onabotulinumtoxinA into hypertonic pelvic floor muscles should be used in conjunction to PFPT in women who do not achieve adequate improvement with PT alone. | 8 (73) |
| When using onabotulinumtoxinA in the pelvic floor, should a standard template injection or a targeted injection at hypertonic muscles be used? Targeted | 9 (82) |
| Should injections of onabotulinumtoxinA be performed unilateral or bilateral? Bilateral | 8 (73) |
| If no improvement with onabotulinumtoxinA after how many sessions do you move on to another therapy? 1-2 | 10 (91) |
| There was a wide variation in the amount of botuliumtoxin per muscle group used from 10 to 50 units per muscle group. Units per muscle group depends on how many muscle groups are being targeted but is somewhere between 10-40 units per muscle group not to exceed 360 units in 3 months. | 8 (73) |
| *Vaginal Muscle Relaxants* |  |
| Vaginal administration of benzodiazepines, such as diazepam, may reduce pelvic floor muscle tone. Although individual response is variable, it is worth a trial given low risk and invasiveness of therapy. | 8 (73) |
| Studies have shown that vaginally administered diazepam results in lower peak serum plasma concentrations, and longer half-life, resulting in fewer side effects. Thus, vaginal administration is preferred over oral administration. | 8 (73) |
| Which of the following vaginal muscle relaxants do you use? Diazepam | 8 (73) |
| *Cognitive Behavioral Therapy* |  |
| 80% of respondents believed that psychological counseling or CBT should be offered at some point in the treatment algorithm. Do you feel patients should be offered CBT or psychological counseling as part of the initial assessment after diagnosis of HTPFD is made, OR should it be a second line option if no improvement is made with PFPT. Second line | 8 (73) |
| *Tender Point Injections (TPI)* |  |
| There was a mixed response on the use of pudendal blocks in HTPFD. Respondents felt that pudendal blocks should only be used if there are signs of pudendal neuralgia or as a diagnostic tool to see if the process is peripheral or sympathetically maintained. Others felt this is to treat a different pathophysiology that HTPFD. Pudendal blocks do not have role in pure high-tone pelvic floor dysfunction. | 8 (73) |
| *Sacral Neuromodulation* |  |
| There was a mixed response to the use of sacral neuromodulation (SNM) for HTPFD. 60% of respondents believed it should be included in the treatment algorithm in some capacity however due to the invasiveness of the procedure it should be reserved as a “last resort” option, after trial of less invasive options. Studies have shown the utility of SNM in patients with chronic pelvic or bladder pain with improvement in pelvic pain and urgency with 80% satisfaction (Marinkovic, 2011). The goal of SNM is to target LUT symptoms as well as pelvic pain. Do you agree with the use of SNM as a 4th line option for HTPFD after other options have been tried? | 9 (82) |

| *Therapies Not to Include* |  |
| --- | --- |
| Home therapies such as stretching, heating pads and yoga should be used as first line therapy for HTPFD. | 9 (82) disagreement |
| At home TENS units can be considered as an alternative to PFPT for treatment of HTPFD. | 10 (91) disagreement |
| *Order of therapy* |  |
| Pelvic floor physical therapy should be first line therapy for high-tone pelvic floor dysfunction. | 11 (100) |
| Vaginal muscle relaxants, TPI, and BTX-A should all be considered second line therapies. | 8 (73) |
| Second line treatment options for HTPFD should include (choose 3)<br>Vaginal muscle relaxants, TPIs | 8 (73) |
| Third line treatment options for HTPFD should include (choose 3) BTX-A | 8 (73) |
| Fourth line treatment options for HTPFD should include (choose 3)<br>Sacral neuromodulation | 8 (73) |
| Based on consensus opinions determined from Questionnaire 1, the treatment algorithm should include PFPT as first line option. | 9 (82) |

Ultimately, the experts reached consensus that HTPFD treatment should be arranged into four tiers based on available data and perceived effectiveness and invasiveness of the therapy. Treatment approaches that did not reach expert consensus were not included in the final treatment algorithm. Experts unanimously agreed that after the diagnosis of HTPFD is made, patients should be referred to PFPT and counseled on home-based options for symptom control. Behavioral interventions include timed voiding, urge suppression, and dietary modifications for bowel and bladder symptoms. Home-based stretching exercises with the goal of pelvic floor relaxation and yoga should be routinely recommended. At the time of diagnosis an evaluation should rule out any extra-pelvic myofascial contributions, such as lower extremity joint or spinal pathology, and if identified, a referral should be made to the appropriate specialists for management in conjunction with the pelvic floor measures.

### Pelvic Floor Physical Therapy (PFPT)

There was unanimous consensus that PFPT should be offered as first-line treatment for HTPFD. Consensus was met that PFPT should be employed for a least 8-12 weeks, noting that patients with a longer symptom history may require a greater number of sessions to appreciate improvement with PT. PFPT is aimed at pelvic floor relaxation, not strengthening, employing techniques such as myofascial release and dry needling. Following the initial treatment, women who improve with PFPT may continue therapy until the symptoms stabilize or resolve. Patients may then be discharged with self-management techniques for symptom maintenance and for the treatment of minor flares. After 4-6 months of stable symptoms, patients may follow up on an as-needed basis. Experts agreed that the goal of PFPT is to facilitate initial improvements and provide sufficient training to allow patients to manage symptoms and minor flares with home-based treatment.

Experts agreed that second-line therapies, such as tender point injection (TPI) and vaginal suppositories, can be added to PFPT in patients whose progress has plateaued without sufficient improvement or who are not initially able to tolerate therapy. Experts indicated techniques such as biofeedback and dry needling may be employed at the discretion of the physical therapist and do not need to be specifically requested by the clinician. If symptoms do not improve with PFPT, then an alternative diagnosis should be considered before progression to second-line options.

#### Second Line Therapies

##### Vaginal Muscle Relaxants

Diazepam was the most prescribed vaginal suppository with the goal of muscle relaxation, however other viable options include baclofen, cyclobenzaprine and tizanidine. Vaginal administration was preferred by the experts over oral administration.

##### Tender Point Injections (TPI)

A myofascial trigger point is a tender nodule within a taught band and is more recently being referred to as tender point with increased tone. (15) Tender points often accompany HTPFD, however not all hypertonic muscles have tender points and thus this therapy is most useful for patients with identifiable tender points on exam. To identify an appropriate site for injection, pelvic floor muscles are palpated perpendicular to its fiber orientation for a taut band. The taut band is then palpated within its fiber direction for the most tender spot that reproduces or refers pain. Injection of local anesthetic is targeted at this site. Consensus was reached that a plain local anesthetic (e.g., 0.25%-0.5% bupivacaine) should be used. The majority of experts did not recommend adding steroid to the injection mixture. This is helpful for patients who cannot tolerate PFPT due to a high level of pain or who have plateaued with PFPT. The goal of the TPIs is to downregulate the guarding reflex and allow for further relaxation of the muscle. While response to TPI can be a good prognostic sign, if a patient does not respond to an initial injection, repeated injections are not warranted. Experts did not advocate isolated pudendal blocks in isolation were useful for treating HTPFD.

##### Cognitive Behavioral Therapy (CBT)

CBT has been shown to make a clinically meaningful improvement in pain and psychosexual function in patients with vestibulodynia and other chronic pain conditions thus experts supported its use in HTPFD. (16) (17)

#### Third-Line Therapy

##### OnabotulinumtoxinA (BTXA)

Experts reached consensus that injection with onabotulinumtoxinA is helpful for women with refractory myofascial pelvic pain and should be the third-line option for women who have been minimally responsive to other therapies. Experts reached consensus that BTXA injections should be targeted at the myofascial sources of hypertonicity or pain, rather than administered in a single template fashion, and that bilateral injection is preferred over unilateral. Repeat injections is not recommended if there is no response to the first injection.

#### Fourth-Line Therapy

##### Sacral Neuromodulation (SNM)

Experts anecdotally reported that SNM for urinary urgency and frequency in the setting of HTPFD is sometimes effective to relieve pain, as well as LUTS. Given the paucity of data, however, experts felt SNM should only be used in patients who have failed other therapies and should not be recommended in the absence of refractory frequency and urgency as SNM used primarily for HTPFD would be an off-label use.

## DISCUSSION

Here, we present a first-of-its-kind treatment algorithm for patients with HTPFD. While the first line option of PFPT easily met consensus among our panel of national experts, later lines of therapy with less evidence were supported based on expert opinion and anecdotal success. Specifics regarding technique and dosing of second- and third-line therapies did not meet consensus due to lack of high-quality evidence and heterogenicity in practice patterns. This highlights areas where further research is needed to help refine this treatment algorithm and improve the care of HTPFD.

While PFPT has the most high-quality data to support its use, there are challenges with its application. Most patients cannot identify an appropriate provider, experience long waits, or undergo inadequate or inappropriate treatment. Inadequately trained therapists may focus on muscle strengthening techniques, which exacerbates HTPFD. (18) Finding a skilled provider is challenging as only 305 specialist therapists in the U.S. are registered on the American Physical Therapy Association website; multiple states have only a few board-certified therapists. Uptake and attendance with prescribed PFPT for HTPFD is poor, between 15-40%. (19) (20) Even when delivered by well-trained personnel, PFPT is only effective for two-thirds of patients. (6) In addition, patients need to be physically and cognitively intact and able to actively participate in such therapies. Multiple barriers to starting and completing PFPT have also been identified, and there are no accepted therapeutic equivalents for patients who cannot access physical therapy, cannot participate in self-care, or fail to respond to this approach. (21)

Despite limited guidance regarding formulation, dose, and administration, intravaginal diazepam, a second-line treatment option in the algorithm, is increasingly being used off-label for HTPFD management. (22) (23) Dosing has been adapted from oral dosing and experts felt a starting dose of 5-10 mg vaginally was appropriate **(Table 2)**. Studies have shown that vaginally administered diazepam results in lower peak serum plasma concentrations, longer half-life, and fewer side effects, supporting the recommendation for vaginal over oral administration. (23) (24) As there are no commercially available vaginal diazepam formulations, and compounded medications are typically not covered by insurance, this treatment method can pose a financial barrier for some patients. As the absorption of oral tablets placed vaginally is poor and therapeutic benefit is reported to be less effective, it was not felt to be a universally acceptable alternative. (25) Until insurance coverage improves for compounded vaginal preparations, this therapy is not a universally accessible option.

**Table 2.** Vaginal Muscle Relaxant Options.

| Medication | Dosing Options |
| --- | --- |
| Diazepam | 5-10 mg as needed up to three times per day<sup>24</sup> |
| Tizanidine | 2 to 4 mg at bedtime* |
| Baclofen | 30mg daily to three times a day* |
| Cyclobenzaprine | 5 to 10mg at bedtime* |

\*Based on expert opinion

Psychosocial factors are well known to play a role in chronic pain, dictating severity and prognosis. Comorbid depression and anxiety among women with chronic pelvic pain are reported as high as 66%. (26) In addition, central sensitization, or the amplification of the central nervous system response to peripheral input, may also occur in patients who experience chronic pain. The combination of these factors highlights how CBT may be an acceptable treatment option for pain due to HTPFD and pain-related depression and anxiety. Although studies of CBT in patients with HTPFD are lacking, strong evidence for its benefits in a wide range of chronic pain disorders and low risk of harm make CBT a reasonable second-line therapy.

While off-label pelvic floor BTXA injection has been used successfully for HTPFD, there is a wide variation in its use and application. Clinically significant reduction in pain score and resting muscle tone are observed across studies despite wide variability in dose, number of injection sites, and method of injection. (27) Experts indicated injections should be bilateral and targeted to hypertonic muscles using 10-40 units per muscle group. However, there remain many unanswered questions regarding optimal injection technique, identification of injection sites and injection amount that needs further investigation with larger randomized controlled trials before definitive recommendations on BTXA can be put forth.

There are no studies specifically evaluating the use of SNM for the treatment of HTPFD. SNM’s therapeutic effect in treating HTPFD is hypothesized to be related to reviving brainstem autoregulation and reset the function of the pelvic floor as evidence by its utility in patients with Fowler’s syndrome. (28) Furthermore, SNM improves pain scores between 35 to 52% in patients with non-IC/BPS chronic pelvic pain, including those with HTPFD. (29) While the expert panel did not feel SNM should be used in patients without urinary or fecal urgency and frequency, data suggest there may be a benefit of SNM for HTPFD alone even in the absence of these complaints; further efforts are needed to evaluate this indication.

Recognizing that the first- and second-line therapies listed in the algorithm are only accessible in resource-rich settings, we specifically asked experts for options in resource-limited settings. Virtual PFPT has increased accessibility in recent years, and experts felt this was an acceptable alternative for patients with no or limited in-person options. The caveat to this is that it must be clearly communicated that therapy should focus on pelvic floor relaxation. Experts also felt progression straight to vaginal muscle relaxants and TPIs was a reasonable option. Overall, experts reiterated their frustration that patients experience numerous barriers to obtaining even these limited options, and improved insurance coverage and access to care is needed to make these therapies more accessible. Sociodemographic inequalities have been highlighted as barriers to pain management access and may be even greater in patients with HTPFD. (30)

The strength of this novel consensus algorithm for the treatment of HTPFD derives from broad experiences of an expert panel with diverse medical specialties and geographic locations, strengthening the generalizability of this algorithm. However, this algorithm is limited in the ability to provide specific guidance on the application of the therapeutics beyond PFPT due to poor quality research. Even retrospective studies on an HTPFD population are limited by no specific ICD10 diagnosis codes for HTPFD. Without wider recognition of this condition, improvements in the diagnostic recognition and therapeutic offerings will continue to stagnate.

With this expert-driven treatment algorithm, patients can now progress through treatment in a systematic fashion. This will give providers more confidence when recommending therapies and patients more confidence that additional options are available if they do not respond or cannot access first-line options. This will also allow for further studies on responses to therapy, improving the quality of data on these treatment decisions. In turn, this algorithm can be revised and updated as research develops in this field.

## Conclusion

In summary, this algorithm was created by a diverse group of experts according to rigorous criteria to generate a consensus treatment pathway for HTPFD supported by available literature. PFPT is unanimously agreed upon to be first-line treatment of this condition. The second-, third-, and fourth-line options are available for patients who do not adequately respond to PFPT. In general, the expert panel reached consensus that research on the treatment of HTPFD has been limited due by diagnostic challenges and poor awareness of this condition. With clear treatment recommendations, providers can guide patients through treatment of HTPFD in a more seamless manner, improving awareness, efficacy, outcomes and satisfaction with care.

## Data Availability

All data produced in the present study are available upon reasonable request to the authors

## Acknowledgments

The study team members acknowledge with gratitude the panelists who completed the Delphi surveys.

## Appendix

### Appendix 1 Statements That Did Not Meet Consensus

Appendix 1- Statements That Did Not Meet Consensus
| Statement | No. (%) Agreement (n=11) |
| --- | --- |
| *Biofeedback* |  |
| Women who continue to have moderate to severe symptoms (i.e. interfering with daily activities or sleep) after completion of pelvic floor physical therapy should be treated with biofeedback. | 4 (36) |
| Biofeedback should be first line for the treatment of HTPFD. | 3 (27) |
| Biofeedback should be used if PFPT has been tried and success as plateaued or not been achieved. | 6 (55) |
| There is a standard protocol that is used for biofeedback? | 2 (18) |
| If pelvic floor EMG biofeedback is available in the clinician's office (i.e. urostym) it can be considered as an alternative to PFPT particularly for patients who cannot access PFPT. | 3 (27) |
| *Vaginal Muscle Relaxants* |  |
| For patients who cannot afford compounded vaginal muscle relaxants, oral muscle relaxants are the best alternative. | 2 (18) |
| For patients who cannot afford compounded vaginal muscle relaxants, oral muscle relaxants placed vaginally are the best alternative. | 3 (27) |
| Vaginal muscle relaxants should be tried before pelvic floor TPI or BTX-A is considered. | 4 (36) |
| Vaginal muscle relaxants should be used in conjunction with PFPT for the treatment of HPTFD. | 4 (36) |
| *BTX-A and TPI* |  |
| BTX-A ideally should be combined with a local anesthetic like Marcaine. The anesthetic portion can be helpful if the HTPFM is due to a sensory pain trigger at the periphery that can be blocked. Thus, the person can learn to relax the muscle once the trigger is removed. | 6 (55) |
| Ideal timing of TPI is injection in the physician's office followed by an immediate PT session within 1 hour. | 3 (27) |
| 60% of respondents agreed that BTX-A should be considered in conjunction with PFPT if there is a plateau in improvement. In this situation BTX-A should be considered based on the recommendation by PFPT. | 7 (64) |
| BTX-A or TPI are more effective than vaginal muscle relaxants and thus should be tried first. | 3 (27) |
| Injection of local anesthetic to hypertonic muscles should be used in conjunction with BTX-A. | 7 (64) |
| Injection of local anesthetic to hypertonic muscles helps treat HTPFD. | 6 (55) |
| How many units of BTX-A should be used per muscle group? | No consensus |
| Pudendal blocks should be used in conjunction with BTX-A injections to the pelvic floor. | 0 (0) |
| To determine a patient's response to BTX-A injections, an initial trial in the office of an injection into the hypertonic muscle should be used. If there is improvement, then BTX-A should be used for subsequent injections. If no improvement alternative therapies should be recommended. | 3 (27) |
| Injection of BTX-A into hypertonic pelvic floor muscles should only be used in patients who are refractory to biofeedback, PFPT and vaginal muscle relaxants. | 6 (55) |
| *Sacral Neuromodulation* |  |
| Sacral neuromodulation should be considered if patient has failed home therapy, biofeedback, pelvic floor physical therapy, vaginal muscle relaxants and TPIs. | 5 (45) |
| Sacral neuromodulation should be considered if patients have failed PFPT. | 4 (36) |
| *Alternative Therapies* |  |
| Women with minimal or no improvement with PFPT should be reassessed and referred for evaluation with a psychiatrist or orthopedist to assess for extra-pelvic sources of pain. | 5 (45) |
| There is evidence to support the use of home therapies such as stretching, heating pads and yoga for the treatment of HTPFD. | 5 (45) |
| At home TENS units have a role in the treatment of HTPFD. | 2 (18) |

## REFERENCES

1. Chronic Pelvic Pain: ACOG Practice Bulletin, Number 218 2020 [135(3):e98–e09. doi:10.1097/AOG.0000000000003716]. Available from: https://www.acog.org/clinical/clinical-guidance/practice-bulletin/articles/2020/03/chronic-pelvic-pain.

2. Mathias SD, Kuppermann M, Liberman RF, Lipschutz RC, Steege JF. Chronic pelvic pain: Prevalence, health-related quality of life, and economic correlates. Obstetrics and gynecology. 1996;87(3):321–7.

3. Ross V, Detterman C, Hallisey A. Myofascial Pelvic Pain: An Overlooked and Treatable Cause of Chronic Pelvic Pain. J Midwifery Womens Health. 2021;66(2):148–60.

4. Westbay LC, Adams W, Kistner M, Brincat C, Bresler L, Yang LC, et al. Clinical Outcomes of a Multidisciplinary Female Chronic Pelvic Pain Program. Female Pelvic Med Reconstr Surg. 2021;27(12):753–8.

5. Voorham-van der Zalm PJ, Lycklama ANGAB, Elzevier HW, Putter H, Pelger RCM. “Diagnostic investigation of the pelvic floor”: a helpful tool in the approach in patients with complaints of micturition, defecation, and/or sexual dysfunction. J Sex Med. 2008;5(4):864–71.

6. Bedaiwy MA, Patterson B, Mahajan S. Prevalence of myofascial chronic pelvic pain and the effectiveness of pelvic floor physical therapy. J Reprod Med. 2013;58(11-12):504–10.

7. Faubion SS, Shuster LT, Bharucha AE. Recognition and management of nonrelaxing pelvic floor dysfunction. Mayo Clin Proc. 2012;87(2):187–93.

8. Meshkat B CS, Gethin G, et. al. Using an e-Delphi technique in achieving consensus across disciplines for developing best practice in day surgery in Ireland. J Hosp Adm. 2014;3(4).

9. Murphy MK, Black NA, Lamping DL, McKee CM, Sanderson CF, Askham J, et al. Consensus development methods, and their use in clinical guideline development. Health Technol Assess. 1998;2(3):i–iv, 1–88.

10. Giannarou L ZE. Using Delphi technique to build consensus in practice. Int J Bus Sci Appl Manag. 2014;9(2):65–82.

11. Messelink B, Benson T, Berghmans B, Bo K, Corcos J, Fowler C, et al. Standardization of terminology of pelvic floor muscle function and dysfunction: report from the pelvic floor clinical assessment group of the International Continence Society. Neurourol Urodyn. 2005;24(4):374–80.

12. Atkins D, Eccles M, Flottorp S, Guyatt GH, Henry D, Hill S, et al. Systems for grading the quality of evidence and the strength of recommendations I: critical appraisal of existing approaches The GRADE Working Group. BMC Health Serv Res. 2004;4(1):38.

13. Hasson F, Keeney S, McKenna H. Research guidelines for the Delphi survey technique. J Adv Nurs. 2000;32(4):1008–15.

14. T S. The Delphi Technique: An Adaptive Research Tool. Br J Occup Ther. 1998;61(4):153–6.

15. Frawley H, Shelly B, Morin M, Bernard S, Bo K, Digesu GA, et al. An International Continence Society (ICS) report on the terminology for pelvic floor muscle assessment. Neurourol Urodyn. 2021;40(5):1217–60.

16. Goldfinger C, Pukall CF, Thibault-Gagnon S, McLean L, Chamberlain S. Effectiveness of Cognitive-Behavioral Therapy and Physical Therapy for Provoked Vestibulodynia: A Randomized Pilot Study. J Sex Med. 2016;13(1):88–94.

17. Urits I, Callan J, Moore WC, Fuller MC, Renschler JS, Fisher P, et al. Cognitive behavioral therapy for the treatment of chronic pelvic pain. Best Pract Res Clin Anaesthesiol. 2020;34(3):409–26.

18. Hoffman D. Central and peripheral pain generators in women with chronic pelvic pain: patient centered assessment and treatment. Curr Rheumatol Rev. 2015;11(2):146–66.

19. Woodburn KL, Tran MC, Casas-Puig V, Ninivaggio CS, Ferrando CA. Compliance With Pelvic Floor Physical Therapy in Patients Diagnosed With High-Tone Pelvic Floor Disorders. Female Pelvic Med Reconstr Surg. 2021;27(2):94–7.

20. Shannon MB, Genereux M, Brincat C, Adams W, Brubaker L, Mueller ER, et al. Attendance at Prescribed Pelvic Floor Physical Therapy in a Diverse, Urban Urogynecology Population. Pm&R. 2018;10(6):601–6.

21. Zoorob D, Higgins M, Swan K, Cummings J, Dominguez S, Carey E. Barriers to Pelvic Floor Physical Therapy Regarding Treatment of High-Tone Pelvic Floor Dysfunction. Female Pelvic Med Re. 2017;23(6):444–8.

22. Crisp CC, Vaccaro CM, Estanol MV, Oakley SH, Kleeman SD, Fellner AN, et al. Intra-vaginal diazepam for high-tone pelvic floor dysfunction: a randomized placebo-controlled trial. Int Urogynecol J. 2013;24(11):1915–23.

23. Stone RH, Abousaud M, Abousaud A, Kobak W. A Systematic Review of Intravaginal Diazepam for the Treatment of Pelvic Floor Hypertonic Disorder. J Clin Pharmacol. 2020;60 Suppl 2:S110–S20.

24. Larish AM, Dickson RR, Kudgus RA, McGovern RM, Reid JM, Hooten WM, et al. Vaginal Diazepam for Nonrelaxing Pelvic Floor Dysfunction: The Pharmacokinetic Profile. J Sex Med. 2019;16(6):763–6.

25. Hussain A, Ahsan F. The vagina as a route for systemic drug delivery. J Control Release. 2005;103(2):301–13.

26. Siqueira-Campos VME, Da Luz RA, de Deus JM, Martinez EZ, Conde DM. Anxiety and depression in women with and without chronic pelvic pain: prevalence and associated factors. J Pain Res. 2019;12:1223–33.

27. Meister MR, Brubaker A, Sutcliffe S, Lowder JL. Effectiveness of Botulinum Toxin for Treatment of Symptomatic Pelvic Floor Myofascial Pain in Women: A Systematic Review and Meta-analysis. Female Pelvic Med Reconstr Surg. 2021;27(1):e152–e60.

28. Dasgupta R, Critchley HD, Dolan RJ, Fowler CJ. Changes in brain activity following sacral neuromodulation for urinary retention. J Urol. 2005;174(6):2268–72.

29. Mahran A, Baaklini G, Hassani D, Abolella HA, Safwat AS, Neudecker M, et al. Sacral neuromodulation treating chronic pelvic pain: a meta-analysis and systematic review of the literature. Int Urogynecol J. 2019;30(7):1023–35.

30. Knoebel RW, Starck JV, Miller P. Treatment Disparities Among the Black Population and Their Influence on the Equitable Management of Chronic Pain. Health Equity. 2021;5(1):596–605.

